# Post-stroke Epilepsy and its Association with Stroke Location – Systematic review and Meta-analysis

**DOI:** 10.1101/2024.04.08.24305465

**Authors:** Sihyeong Park, Yasar T. Esengul, Khaled Gharaibeh, Sidra Saleem, Ajaz Sheikh

**Author notes:** Correspondence to: Sihyeong Park, 200 1^st^ St SW, Rochester, MN 55901.

## Abstract

**Objective:** There have been several meta-analytic studies that investigated potential risk factors for post-stroke epilepsy (PSE)^1,2^. Cortical location is known to be associated with increased risk of PSE. However, relationship between the involvement of specific lobe with stroke and PSE has not been evaluated in a meta-analysis due to unavailability of relevant data. To the best of our knowledge, this is the first meta-analysis on this subject matter.

**Methods:** The PubMed and Embase databases were comprehensively assessed and searched for studies that evaluated the association between the involvement of different lobes with PSE. Studies that evaluated post-stroke epilepsy or recurrent post-stroke seizure were included, and acute symptomatic seizure was excluded. Following search strategy was used, “post-stroke” OR “stroke” OR “intracerebral hemorrhage” AND “epilepsy” OR “seizure” AND “lobe” OR “territory” OR “location”. Joanna Briggs Institute appraisal tool was utilized to perform quality assessment of the included studies. A random effects model was used to estimate pooled odds ratio of the selected outcomes with 95 percent confidence intervals (CIs), and the study heterogeneity was analyzed.

**Results:** Nine studies^3-11^ were identified with 1328 stroke patients. 9/9 studies showed association of frontal lobe involvement with higher likelihood of PSE (OR=2.10, 95% CI=1.37-3.21, p-value=0.0007). All studies showed association of parietal stroke with increased likelihood of PSE (OR=2.85, 95% CI=1.93-4.22, p-value<0.00001). Previous temporal lobe stroke was associated with increased risk of PSE (OR=3.55, 95% CI=1.95-6.45, p-value<0.00001). In contrast, association between occipital lobe involvement and PSE was not statistically significant (OR=1.37, 95% CI=0.95-1.96, p-value<0.09), and this remained true when subgroup analysis was performed.

**Discussion:** Our meta-analysis shows that PSE is associated with frontal, parietal, and temporal involvement of previous stroke. There was no statistically significant association between occipital lobe involvement and PSE. Further high quality studies are needed to investigate if stroke in one lobe/territory can be more epileptogenic than those in others.

**Key Points:**

- Cerebrovascular disease is the most common etiology of newly diagnosed epilepsy
- Prior stroke involving the frontal, temporal, or parietal lobe is associated with increased risk of post-stroke epilepsy
- Occipital lobe stroke is not associated with increased risk of post-stroke epilepsy, regardless of the age group.

## 1. Introduction

An epidemiological study that included all age group reported that cerebrovascular disease was the most common etiology in patients with defined cause of newly diagnosed epilepsy^12^. With the ageing population, the importance of new-onset epilepsy in the elderly has become more significant. A number of previous literature reported cerebrovascular disease as the most common etiology of the late-onset epilepsy. A large epidemiological study reported stroke incidence of 13.7 million and stroke prevalence of 80.1 million in 2016^13^.

There have been a few different definitions of post-stroke epilepsy (PSE) over the years^14^. Previous literature has steadily defined late-onset seizures occurring after two weeks or more from stroke, which increases the risk of PSE. The incidence of PSE was reported to be 7% in a meta-analysis that included 102008 stroke patients^15^. There have been a few meta-analytic studies that reviewed the risk factors for PSE^1,2^. Cortical location is known to be associated with increased risk of PSE. However, relationship between the involvement of specific lobe with stroke and PSE has not been evaluated in a meta-analysis due to unavailability of relevant data^1^, although there has been some investigations that demonstrated association of parietal lobe infarction with the increased risk of PSE^8^. To the best of our knowledge, this is the first meta-analysis on this subject matter.

## 2. Methods

### 2.1. Search strategy and selection criteria

The PubMed and Embase databases were comprehensively assessed and searched from their inception to July 2023. Studies that evaluated the association between the involvement of different lobes with PSE were identified. Following search strategy was used, “post-stroke” OR “stroke” OR “intracerebral hemorrhage” AND “epilepsy” OR “seizure” AND “lobe” OR “territory” OR “location”. The full search strategy is outlined in the Supplementary Material (Supp 1). Results were reported according to the Preferred Reporting Items for Systematic Reviews and Meta-Analyses (PRISMA) guidelines. Two authors (S.P. and Y.T.E.) independently performed the search, screened titles and abstracts, and assessed eligibility. For included studies, S.P. collected the data from each study. Following data were extracted: first author name, year of publication, study design, total sample size, number of patients with a history of infarct with involvement of a given lobe and infarct sparing that lobe, and number of patients with PSE in each group with lobar involvement. The number of the patients were directly obtained or derived from the available information in the manuscripts of the included studies. With a study that specified concurrent involvement of different lobes (Kopyta 2021), e.g. frontal only vs frontal, temporal, and parietal, the cases with frontal lobe involvement were added, and the cases with no involvement of frontal lobe were added to evaluate for the association of frontal lobe infarcts and poststroke epilepsy. Same strategy was utilized to derive the number of the cases with infarcts affecting the other lobes.

Inclusion criteria were as follows: (1) studies that evaluated post-stroke epilepsy or recurrent post-stroke seizure and (2) original articles. Following studies were excluded: (1) studies that investigated acute symptomatic seizure, (2) studies that did not clarify the chronicity of the seizure onset after stroke, (3) studies with insufficient information regarding anatomic location of the stroke, (4) irrelevant studies, (5) animal or in vitro studies, (6) case reports, conference abstracts, or reviews, and (7) studies in language other than English.

### 2.2 Quality assessment

Joanna Briggs Institute (JBI) appraisal tool was utilized to perform quality assessment of the included studies.

### 2.3. Statistical analysis

Given the variability in study design and the patient population in the studies, random effects model was assumed and Mantel-Haenszel method was utilized to estimate the pooled odds ratio of the selected outcomes with 95 percent confidence intervals (CIs). Study heterogeneity was analyzed with I^2^ statistic. For the I^2^ statistic, 25% to 50%, 50% to 75%, and 75% were considered low, moderate, and high heterogeneity, respectively. Data analysis was conducted using Review Manager 5.3 and R 4.1.1.

## 3. Results

### 3.1. Study selection

Our search revealed 1952 studies through PubMED, and 2664 studies were identified through EMBASE. Of those studies, 852 articles were duplicated and removed from the screening process. After excluding the articles as outlined in Methods, 9 studies were included in the qualitative and quantitative analysis (Figure 1).

**Figure 1.**
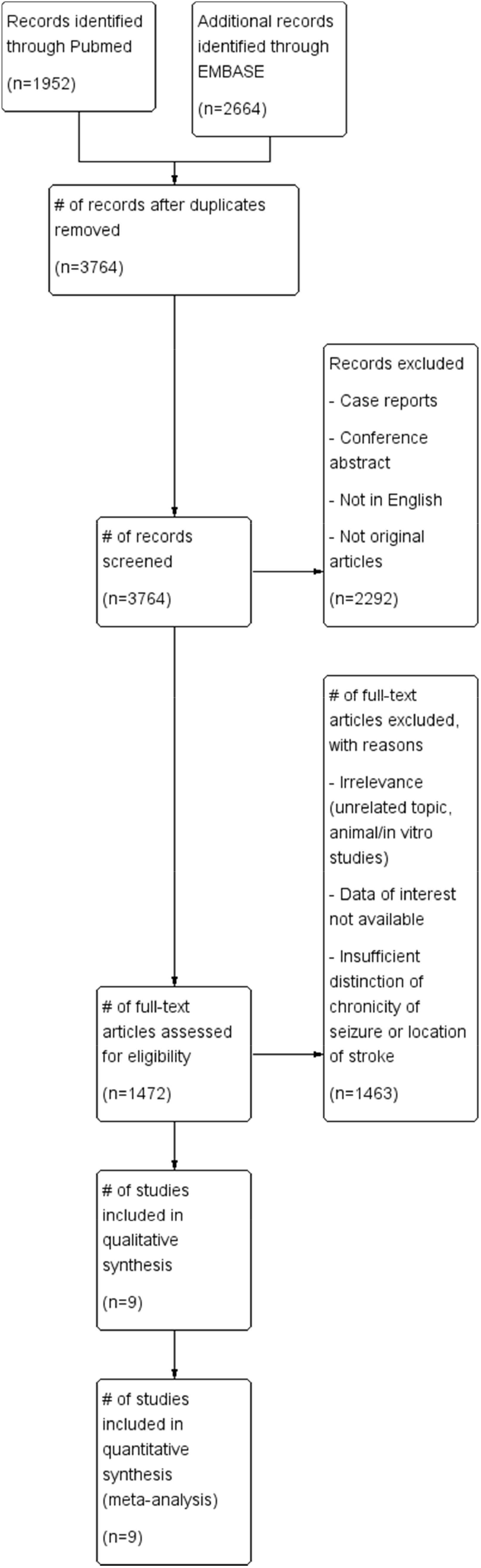
A flow diagram illustrating the study selection process for the review.

### 3.2. Study characteristics

Table 1 illustrates the characteristics of these 9 studies^3-11^, which included a total of 1328 patients with history of stroke. Four of the studies were performed with pediatric population, and the other five included adult population. All the studies were retrospective and observational. The mean age was 56 years, and males represented 57% of total patients. Among 1328 included patients, 255 had PSE (19%). Quality assessment of the included studies with JBI appraisal tool revealed average score of 9.6, with range of 7-11 (Table 2).

**Table 1.** Characteristics of the included studies. d, days; IQR, interquartile range; mo, months; yrs, years.

| Study | Year | Country | Study design | Population | Stroke type | Age | Total number | Male(%) | PSE prevalence (%) |
| --- | --- | --- | --- | --- | --- | --- | --- | --- | --- |
| Vojcek | 2022 | Hungary | Retrospective | Pediatric | Hemorrhagic | Median 60 mo (IQR: 35-88) | 55 | 34 (62) | 9 (16) |
| Ouerdiene | 2021 | Tunisia | Retrospective | Adult | Ischemic | Mean 55.1 yrs | 52 | 39 (75) | 31 (60) |
| Lopez | 2021 | Chile | Retrospective | Pediatric | Ischemic | Median 2 d (IQR 1-5.6) | 33 | 21 (64) | 9 (27) |
| Kopyta | 2021 | Poland | Retrospective | Pediatric | Ischemic | Mean 8.4 ± 5.5 yrs | 75 | 43 (57) | 11 (15) |
| Yamada | 2020 | Japan | Prospective | Adult | Ischemic/Hemorrhagic | Mean 68.2 ± 11.1 yrs | 436 | 257 (58.9) | 26 (6) |
| Takase | 2020 | Japan | Retrospective | Adult | Ischemic | Median case 79.0 (IQR 71 – 86); control 78.5 (IQR 74 – 82) | 93 | 47 (51) | 43 (46) |
| Jia | 2020 | China | Retrospective | Adult | Ischemic | Mean 62.93 ± 8.82 yrs | 209 | 117 (55.98) | 105 (50) |
| Laugesaar | 2018 | Estonia | Retrospective | Pediatric | Ischemic/Hemorrhagic | Median 50 mo (1 mo - 18.4yrs) | 73 | 39 (53) | 21 (29) |
| Keller | 2015 | Germany | Retrospective | Adult | Ischemic | Mean 66.3 ± 12.9 yrs | 302 | 161 (53.3) | 42(13.9) |

**Table 2.** Quality assessment of the studies. *JBI: Joanna Briggs Institute appraisal tool score*.

| ID | Study | JB I (from 11) |
| --- | --- | --- |
| 1 | Vojcek (2022) | 10 |
| 2 | Ouerdiene (2021) | 8 |
| 3 | Lopez (2021) | 7 |
| 4 | Kopyta (2021) | 8 |
| 5 | Yamada (2020) | 10 |
| 6 | Takase (2020) | 11 |
| 7 | Jia (2020) | 11 |
| 8 | Laugesaar (2018) | 11 |
| 9 | Keller (2015) | 11 |

### 3.3. Location of stroke and risk of PSE

All nine studies described the association of frontal lobe stroke and PSE. Prior infarcts with frontal lobe involvement was associated with increased risk of PSE (OR=2.10, 95% CI=1.37-3.21, p-value=0.0007) (Figure 2). I^2^ value was 35%. Subgroup analysis was performed based on the age group. Analysis of the adult subgroup continued to show statistically significant associated between frontal lobe infarct and PSE (OR=2.11, 95% CI=1.43-3.10, p-value=0.0002; I^2^=16%). On contrary, there was no significant association of such in the pediatric group, with higher heterogeneity (OR=2.04, 95% CI=0.51-8.15, p-value=0.31; I^2^ = 61%).

**Figure 2.**
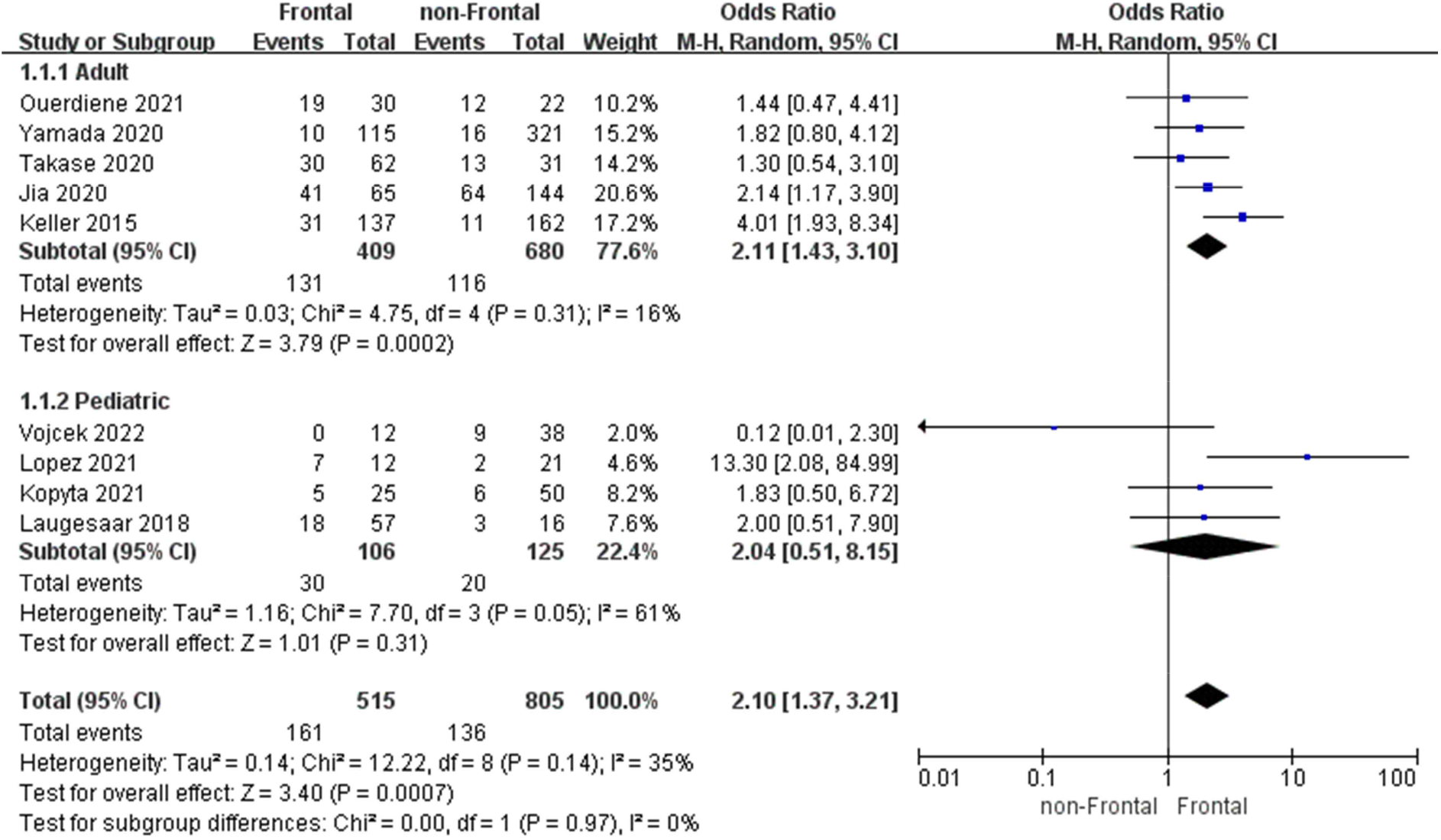
Forest plot illustrating meta-analysis on frontal lobe stroke and PSE.

All included studies showed association of parietal stroke with increased likelihood of PSE (OR=2.85, 95% CI=1.93-4.22, p-value<0.00001). I^2^ value was 24%. Subgroup analysis was performed based on the age group. Risk of PSE was higher in both adult and pediatric subgroups (OR=2.50, 95% CI=1.68-3.70, p-value<0.00001; I^2^=15% and OR=4.15, 95% CI=1.71-10.08, p-value<0.002; I^2^=26%, respectively) (Figure 3).

**Figure 3.**
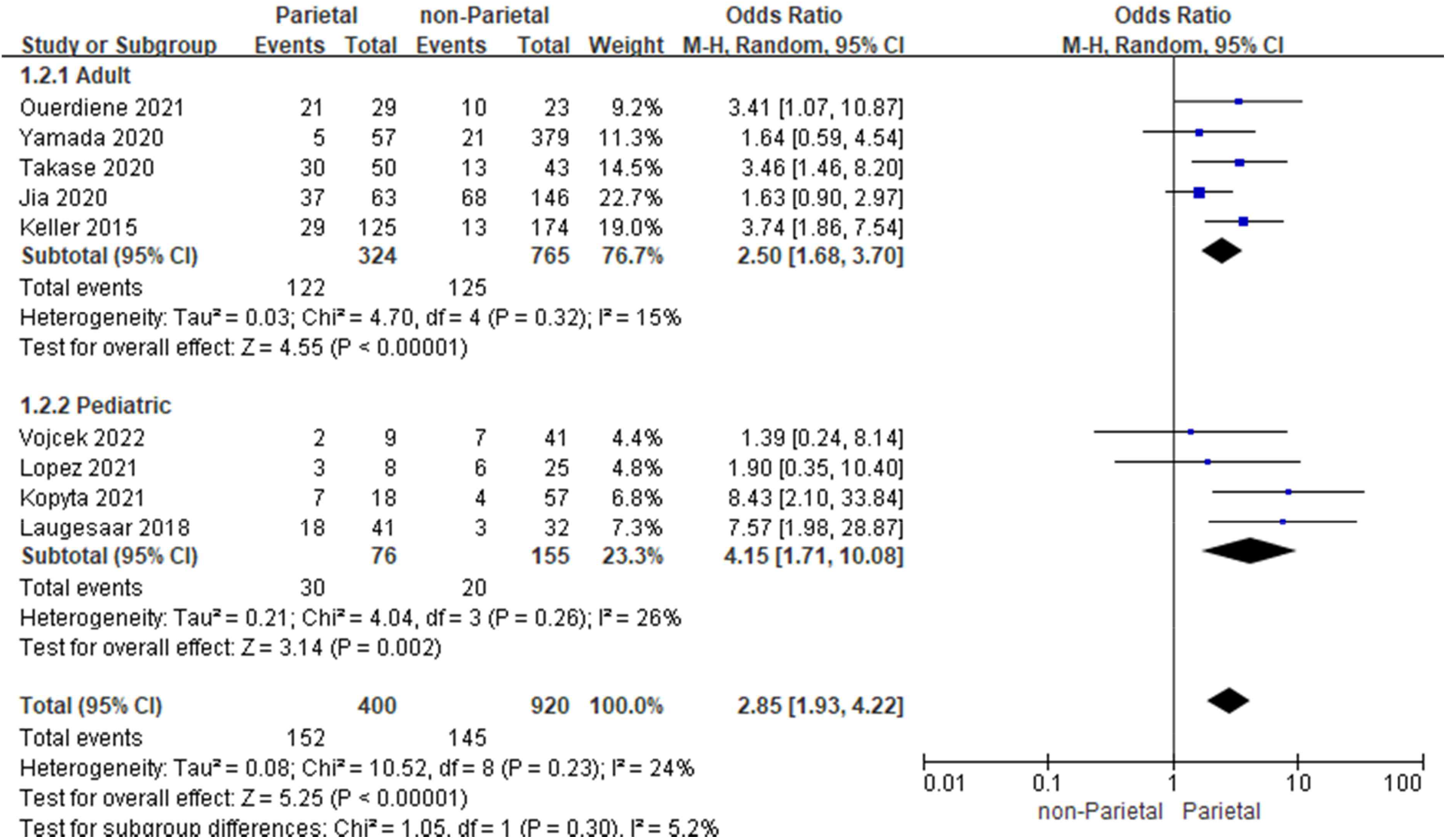
Forest plot illustrating meta-analysis on parietal lobe stroke and PSE.

Previous temporal lobe stroke was associated with increased risk of PSE (OR=3.55, 95% CI=1.95-6.45, p-value<0.00001). I^2^ value was 65%. Risk of PSE in the adult subgroup remained high (OR=2.87, 95% CI=1.67-4.95, p-value=0.0001; I^2^=50%), and in the pediatric subgroup (OR=5.51, 95% CI=1.19-25.45, p-value=0.03; I^2^=75%), although there was a significant heterogeneity in the latter (Figure 4).

**Figure 4.**
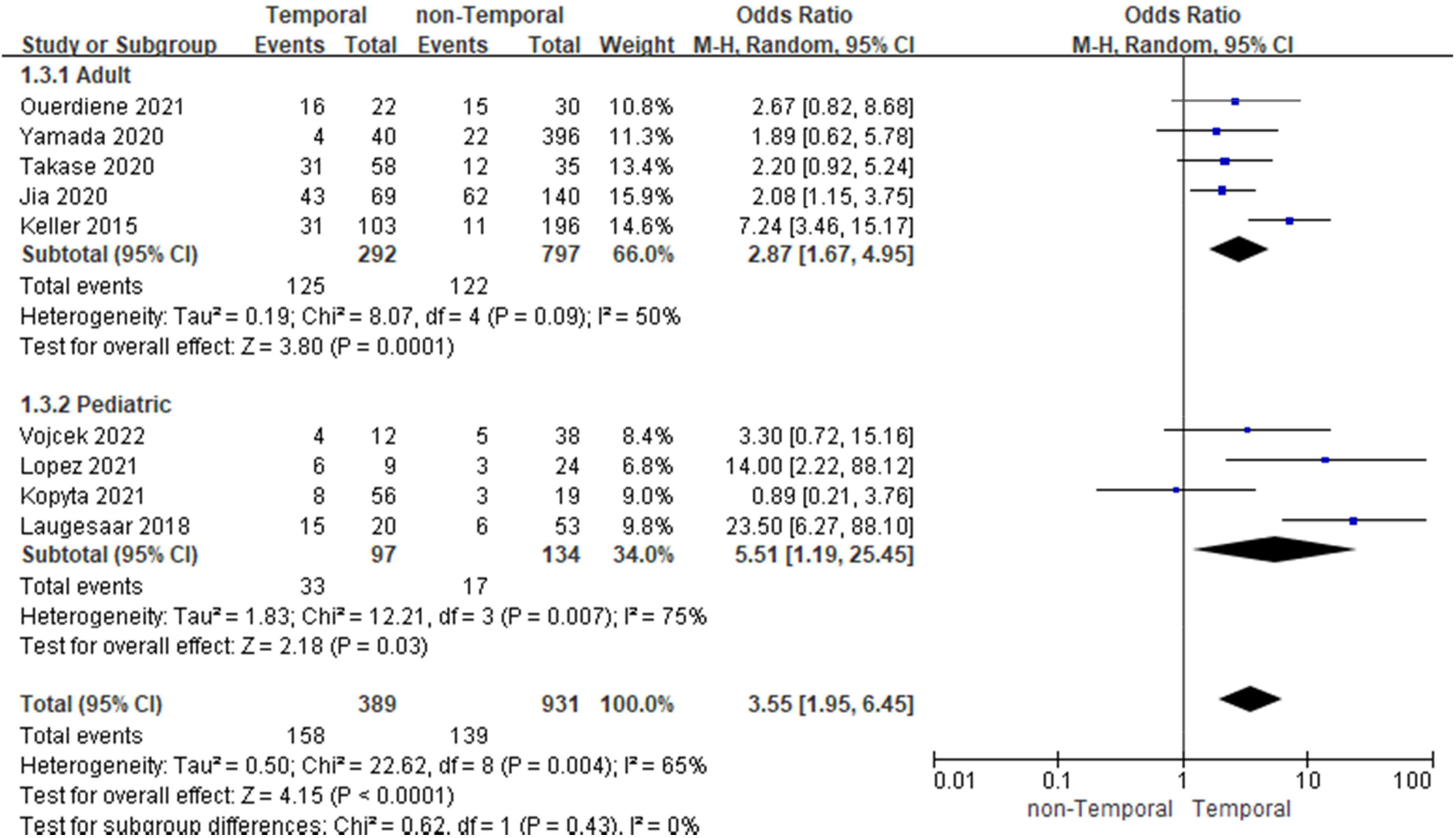
Forest plot illustrating meta-analysis on temporal lobe stroke and PSE.

In contrast, association between occipital lobe involvement and PSE was not statistically significant (OR=1.37, 95% CI=0.95-1.96, p-value=0.09). I^2^ value was 0%. Neither the adult group (OR=1.44, 95% CI=0.98-2.09, p-value=0.06; I^2^=0), nor the pediatric group (OR=0.77, 95% CI=0.22-2.76, p-value=0.69; I^2^=0) showed statistical significance in subgroup analysis (Figure 5).

**Figure 5.**
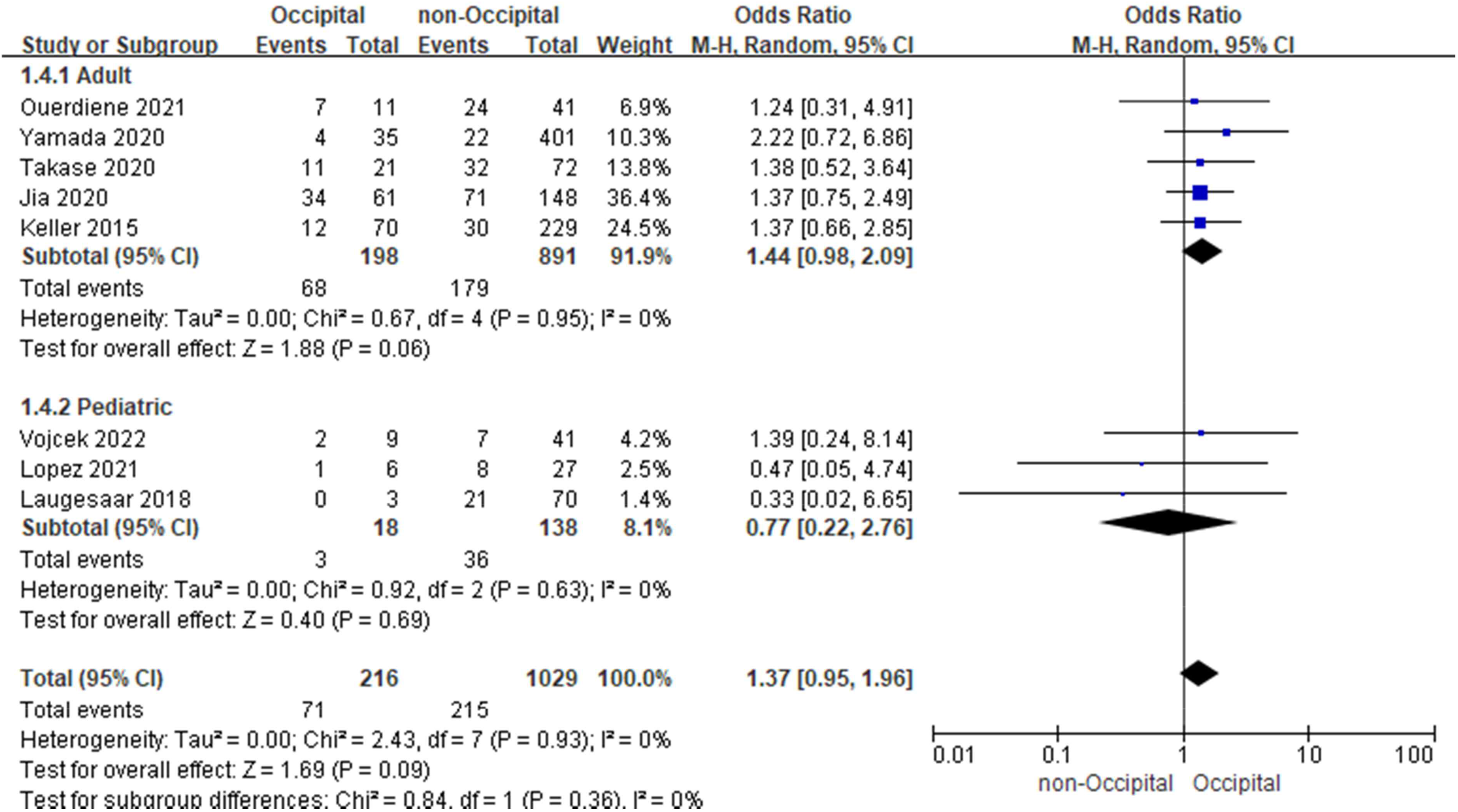
Forest plot illustrating meta-analysis on occipital lobe stroke and PSE.

### 3.3. Risk of bias in studies

Funnel plots of the studies did not reveal significant asymmetry (Figure 6), although the number of the studies was not sufficient to demonstrate a true asymmetry if one existed.

**Figure 6.**
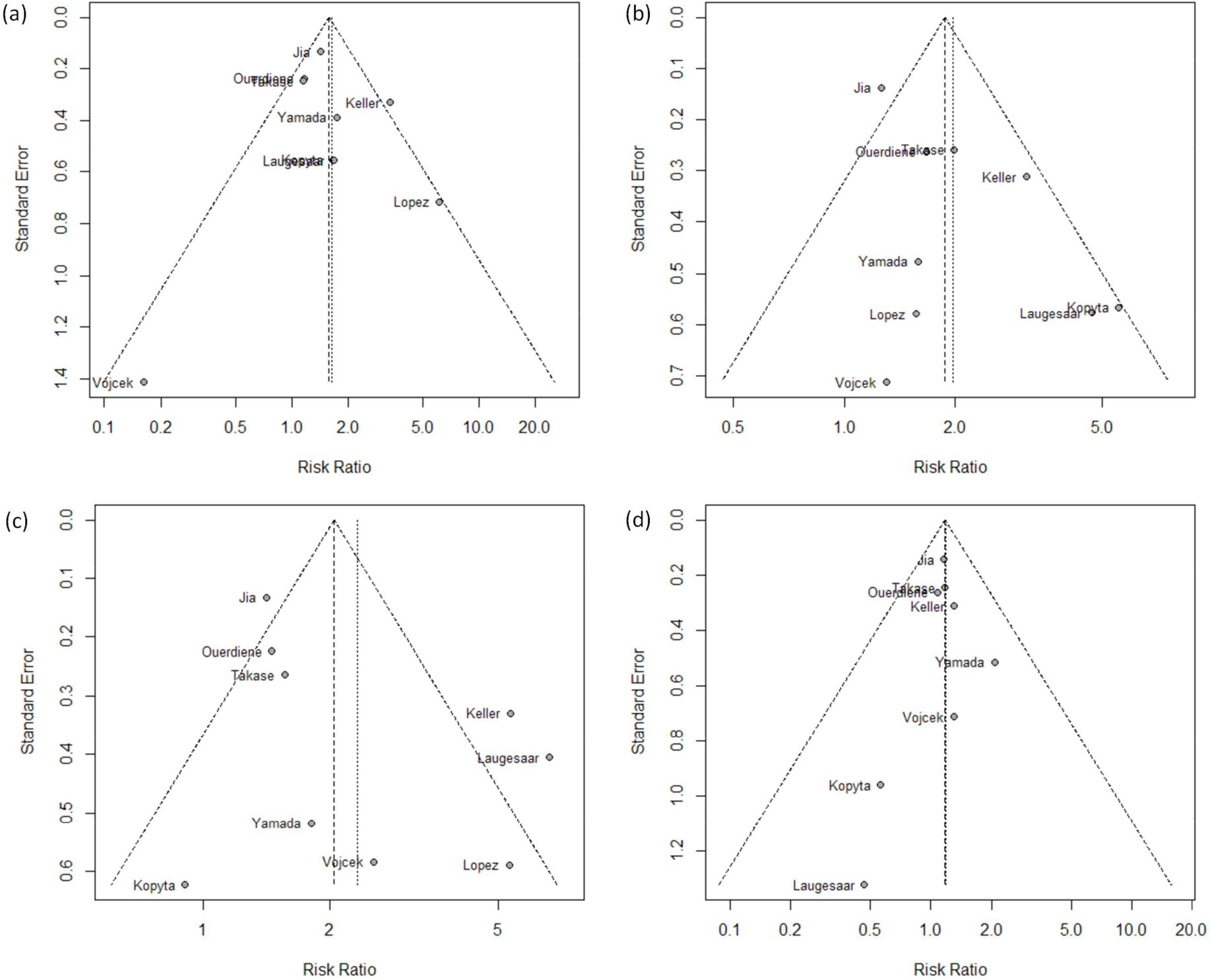
Funnel plots of the studies. (a) frontal lobe, (b) parietal lobe, (c) temporal lobe, and (d) occipital lobe infarcts.

## 4. Discussion

To our knowledge, this is the first meta-analysis to investigate the association between the anatomic location of the ischemic infarct and the risk of PSE. Previous frontal lobe infarct was associated with PSE in adults, but the association was not significant in pediatric population. Parietal lobe infarct and temporal lobe infarct were associated with PSE. There was no such association was observed in patients with occipital lobe infarcts.

There has been growing body of literature regarding the pathophysiology of poststroke epilepsy. The role of astrocytic proliferation and glutamate-releasing channel in excitotoxic complications, such as seizure, after stroke has been postulated in an animal study^16^. Another in vitro study with primary hippocampal culture demonstrated that the neurons survived from stroke-like injury showed spontaneous, recurrent epileptiform discharges^17^. Genetic polymorphism associated with PSE has been reported in previous human studies^18,19^.

Interestingly, we found no association between previous occipital lobe infarcts and PSE. It can be postulated that the occipital lobe is resistant to epileptogenesis. Another explanation is that the occipital lobe has fewer or less susceptible ictal networks with other cortical or subcortical regions compared to other lobes. Such postulation is supported by studies that described lack of correlation between occipital lobe involvement of glioma and epilepsy^20^. In addition, it is possible that the rate of reported seizures in patients with occipital lobe infarcts was falsely low due to the relative subtlety of their semiology compared to seizures originating from other cortical areas.

Limitations exist in our study. There was only one prospective study and the rest were retrospective in nature. Also, the prevalence of PSE in the pooled population was lower than that of a previous study^21^. In addition, not all of the studies confirmed seizure onset zones, which may be discordant with the locations of the previous infarcts, although diagnosis of epilepsy does not require this information.

Our analysis was limited by how variables were reported by the included studies. Specifically, only one study^8^ reported the median proportion of each lobe involved by infarction and median infarction volume in each lobe. Although other studies reported other variables, these were not reported separately for stroke involving each lobe, which is required to perform meta-regression analysis.

Analysis showed low to moderate heterogeneity. In general, higher degree of heterogeneity was observed with studies that included pediatric population. Studies that classified the pre-existing infarcts into vascular territories, rather than lobes, were not included, and this may have contributed to the bias and heterogeneity.

Current literature recommends against initiating antiseizure medications in patients who suffered stroke, partly due to the low incidence of PSE. More risk factors in stroke patients for developing PSE are being identified and investigated, including location of the stroke. Further high-quality prospective studies and basic research are needed to deepen the understanding on the mechanism of epileptogenesis of different stroke locations and guide therapeutic approaches for these patients.

## 5. Conclusion

Our meta-analysis shows that PSE is associated with frontal, parietal, and temporal involvement of previous stroke. However, there was no statistically significant association between occipital lobe involvement and PSE. Further high quality studies are needed to investigate if stroke in certain lobe/territory can be more epileptogenic than those in others.

## Data Availability

All data produced in the present study are available upon reasonable request to the authors

## Consent for Publication

Not applicable.

## Author Contributions

S.P. designed the study protocol. S.P. and Y.T.E. performed the systematic review. S.P. carried out the statistical analyses. S.P. drafted the manuscript. All authors critically revised the manuscript.

## Declaration of Generative AI and AI-assisted technologies in the writing process

There is nothing to disclose.

