## Supplementary Material 1 for "Post-stroke Epilepsy and its Association with Stroke Location – Systematic review and Meta-analysis"

**Search strategy for Pubmed database**

post-stroke OR stroke OR intracerebral hemorrhage OR infarct* OR subarachnoid hemorrhage OR SAH OR Cerebrovascular disease*[Title/Abstract]

AND

(((epilepsy[MeSH]) OR epileps*[Title/Abstract]) OR seizure[MeSH]) OR seizure*[Title/Abstract]

AND

"lobe"[tw] OR "lobes"[tw] OR "territory"[tw] OR "territories"[tw] OR “location”[tw] OR “locations”

**Search strategy for EMBASE database**

‘post-stroke’:ab,ti OR ‘stroke’:ab,ti OR ‘intracerebral hemorrhage’:ab,ti OR ‘infarct’:ab,ti OR ‘infarcts’:ab,ti OR ‘subarachnoid hemorrhage’:ab,ti OR ‘SAH’:ab,ti OR ‘Cerebrovascular disease’:ab,ti

AND

‘epilepsy’/exp OR ‘seizure’/exp OR ‘epilepsy’:ab,ti OR ‘seizure’:ab,ti OR ‘seizures’:ab,ti

AND

‘lobe’:ab,ti OR ‘lobes’:ab,ti OR ‘territory’:ab,ti OR ‘territories’:ab,ti OR ‘location’:ab,ti OR ‘locations’:ab,ti
